# ODACH: A One-shot Distributed Algorithm for Cox model with Heterogeneous Multi-center Data

**DOI:** 10.1101/2021.04.18.21255694

**Authors:** Chongliang Luo, Rui Duan, Yong Chen

## Abstract

**Objective:** We developed and evaluated a privacy-preserving One-shot Distributed Algorithm for Cox model to analyze multi-center time-to-event data without sharing patient-level information across sites, while accounting for heterogeneity across sites by allowing site-specific baseline hazard functions and feature distributions.

**Materials and Methods:** We constructed a surrogate likelihood function to approximate the Cox log partial likelihood function which is stratified by site, using patient-level data from a single site and aggregated information from other sites. The ODAC estimator was obtained by maximizing the surrogate likelihood function. We evaluated and compare the performance of ODACH with meta-analysis by extensive numerical studies.

**Results:** The simulation study showed that ODACH provided estimates close to the pooled estimator, which is obtained by directly analyzing patient-level data from all sites via a stratified Cox model. The relative bias was <1% across all scenarios. As a comparison, the meta-analysis estimator, which was obtained by the inverse variance weighted average of the site-specific estimates, had substantial bias when the event rate is <5%, with the relative bias reaching 12% when the event rate is 1%.

**Conclusions:** ODACH is a privacy-preserving and communication-efficient method for analyzing multi-center time-to-event data, which allows the baseline hazard functions as well as the distribution of covariate variables to vary across sites. It provides estimates that is close to the pooled estimator and substantially outperforms the meta-analysis estimator when the event is rare. It is thus extremely suitable for studying rare events with heterogeneous baseline hazards across sites in a distributed manner.

## INTRODUCTION

Real-world data (RWD) such as electronic health records (EHRs) and medical claims are drawing more attention in generating real-world evidence to support healthcare decision making [1-3]. The past decade has seen an increasing number of clinical research networks, curating and using various EHRs and medical claims data. For example, the Observational Health Data Sciences and Informatics (OHDSI) network is an international network that cover more than half a billion patient records in a common data model (CDM) [5]. Another example is the national Patient-Centered Clinical Research Network (PCORnet), a network covering more than 100 million patients in the United States [6]. These large data consortia provide platforms and tools to integrate heterogenous RWD from a diverse range of healthcare organizations. Multicenter analyses using RWD from these clinical research networks have been increasing rapidly in large part because they can improve the generalizability of the study results due to increased sample size.

Despite the benefits of multicenter analyses, two major challenges exist for multi-site data integration. First, the direct sharing of patient-level data across institutions may be prohibited, as the individual patient-level data are protected by privacy regulations such as the Health Insurance Portability and Accountability Act (HIPAA) or General Data Protection Regulation (GDPR). Hence, many research networks, such as OHDSI, adopt a federated model in which the patient-level data are stored at local institutions and only aggregated information are shared [6-8]. Second, data from different sites are often heterogeneous due to different patient characteristics, data quality, and other unobserved site-level features. Assuming a common statistical model across all sites may result in biased estimation and poor prediction.

Survival analyses are frequently used in biomedical research to model the time to a specific event of interest. To conduct multi-center survival analyses without sharing patient-level data, a common and convenient approach is the meta-analysis, e.g. a weighted average of the local estimates from each site. However, when the outcomes or exposures are rare, or the sample sizes are small in some sites, the accuracy of the meta-analysis may be poor [19,29]. Other distributed algorithms were developed to obtain more accurate results. For example, WebDISCO (a web service for distributed Cox model learning) [14] were developed for conducting logistic regression and Cox regression respectively. Despite the fact that these algorithms can provide identical results to the analysis using the pooled individual-level data (lossless), they are communication intensive due to the iterative nature of the algorithms which requires multiple rounds of communications across site. To balance the communication efficiency and estimation accuracy, Shu et al. [30] proposed a lossless one-shot algorithm for stratified Cox model which can only include one binary covariate in the model. Wang et al. [31] proposed a divide and conquer approach, which aims to reduce the computational challenge when the sample size is extremely large. In addition, Duan et al. [29] proposed a One-shot Distributed Algorithm for Cox model (ODAC) based on the surrogate likelihood approach which relies on patient-level data from a single site and aggregated data from other sites. The algorithm only requires aggregated data twice from other sites but obtains close estimation as if using patient-level data from all sites.

Reviewing the mentioned approaches, most of them are based on the Cox proportional hazards model, but few of them take the data heterogeneity across sites into account. Specifically, in the multi-center survival analysis, the distribution of covariates is likely to be different across sites as patients might come from different sub-populations. In addition, the baseline hazard functions might also be heterogenous across sites. Assuming a Cox model with a common baseline hazard function may lead to biased results. In this article, we propose a distributed algorithm which account for site-level heterogeneities on covariate distributions and baseline hazard functions, namely the One-shot Distributed Algorithm for Cox model with Heterogeneity (ODACH). Compared to the previous work of ODAC that assumes common baseline hazard function across sites, the assumption of heterogeneous baseline hazard functions in ODACH is more flexible and practical in real-world data. Moreover, the constructed surrogate likelihood does not require one extra round of communication regarding the risk set in each site as in ODAC. As a result, ODACH is more communication efficient over ODAC. We show that our proposed algorithm is one-shot and achieves high accuracy (small bias) in a simulation study.

## METHODS

### Stratified Cox Proportional Hazard Model

To account for site-specific baseline hazards, we introduce the stratified Cox model for time-to-event outcome. Suppose we have study subjects from *K* different clinical sites and denote *n*_*j*_ to be the number of subjects in the *j-th* site. Denote the total number of subjects to be 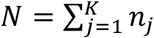. For the *i*-th subject in the *j*-th site, we observe {*T*_*ij*_, *δ*_*ij*_, *x*_*ij*_}, where *T*_*ij*_ is the observed time to event, *x*_*ij*_ is a p-dimensional covariate variable, and *δ*_*i*_ = 0 indicates censoring and *δ*_*i*_ = 1 indicates an event. Assume that the *j*-th site has its own baseline hazard function *λ*_*j*_(*t*), the Cox proportional hazard model assumes that the hazard of the *i*-th subject in the *j*-th site having the event at time *t* follows

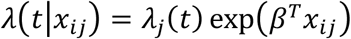

where *β* contains the log hazard ratios. We assume that *β* is the same across all sites, i.e. common effects of the covariates on the time-to-event across sites. For the *j*-th site, the log Cox partial likelihood function is

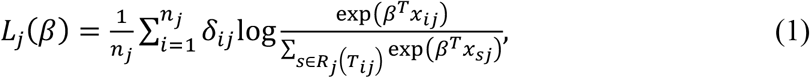

where *R*_*j*_(*t*) is the risk set in site *j* at time *t* defined as *R*_*j*_(*t*) = {*i*; *T*_*ij*_ ≥ *t*}, which contains all the subjects in site *j* that have not experienced an event or been censored at time *t*. Notice that the partial likelihood does not involve the baseline hazard *λ*_*j*_(*t*). To deal with the heterogeneity of baseline hazard functions, we can construct a stratified log Cox partial likelihood function

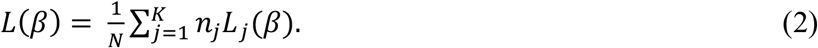

The common effect *β* can be estimated by maximizing the stratified function (1), i.e. 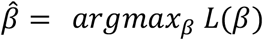. We call it the pooled estimator, as it requires all the data to be pooled together.

### A One-Shot Distributed Algorithm for Cox Model with Heterogeneity

In practice, the patient-level data are often difficult to be transferred across sites. Hence the pooled estimate 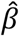 could be hard to obtain. Inspire by the surrogate likelihood approach developed in [9,13,20], we aim to construct a proxy of the stratified Cox partial likelihood function (1), which we call a surrogate likelihood function, using only summary-level information from other sites. Define 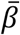 to be an initial value that is close to the true value of the parameter β, and we only have access to the patient-level data of a leading site (e.g. the first site), we propose to construct the following surrogate likelihood function

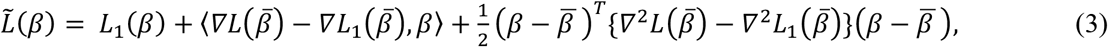

where *L*_1_(*β*) is the local likelihood function defined in (1), *▽* and *▽*^2^ denote the first and second order gradients of a function (explicit forms of 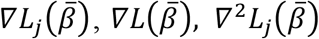 and 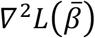 can be found in the Appendix). Intuitively, the above surrogate likelihood function (3) modifies the local likelihood function *L*_1_(*β*) to approximate the stratified likelihood (2), with the modification being the first and second-order terms, i.e. 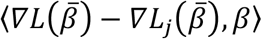 and 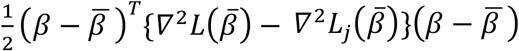. By sharing the second-order gradients, our method allows each site to have different covariate distributions. The surrogate estimator is obtained by maximizing the surrogate likelihood (3), i.e. 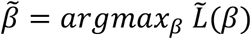.

In the surrogate likelihood function (3), 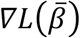 and 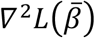 can be calculated distributively by 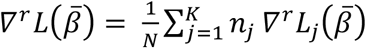, for *r* = 1, 2. Since 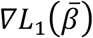 and 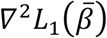 are available from the leading site, it only requires other collaborative sites to calculate and transfer 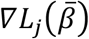 and 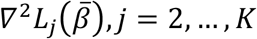. These gradients are all aggregated information, hence patient-level information is protected. Regarding the initial value 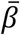, we recommend to use the inverse variance weighted average of the estimates obtained by fitting a Cox model at each site, i.e.,

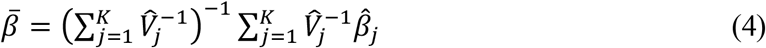

where 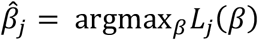 is estimator of Cox model fitted on data in the *j*-th site, and 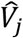 is the estimated vatiance of 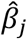. We summarize the ODACH algorithm below and also provide a schematic illutration in Figure 1. Notice in the above derivation, we assume the first site is the leading site when constructing the surrogate likelihood. In practice if all the sites can serve as the leading site, we recommend using the largest site as the leading site. Alternatively, after the derivatives 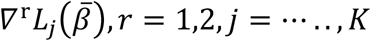 have been shared across sites, each site can serve as the leading site and obtain its own surrogate estimate. These surrogate estimates can be further synthesized to obtain more accurate estimation.

**Figure 1:**
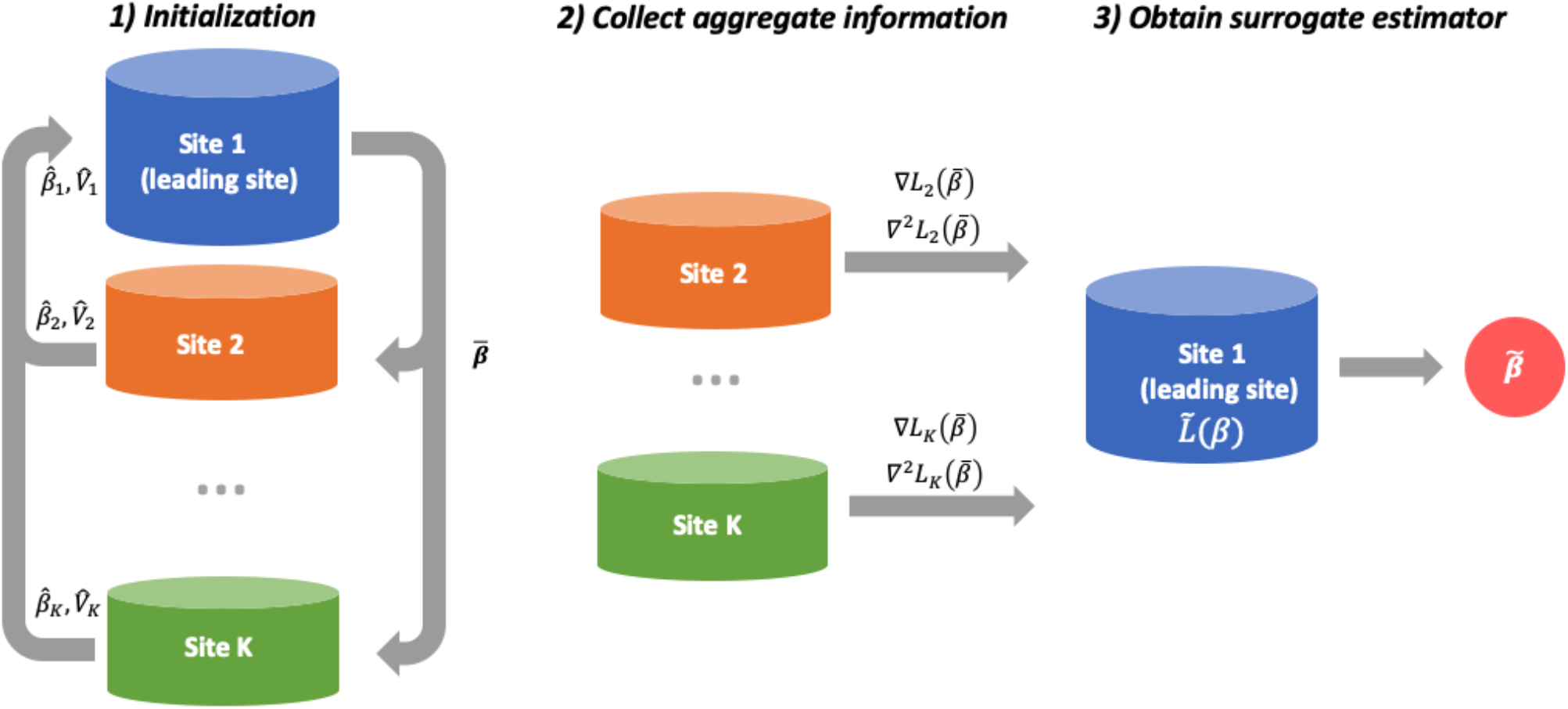
Schematic illustration of the ODACH algorithm. The first step is initialization where each site reports the local estimation of log hazard ratio 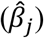, and the corresponding variance estimate 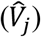. The leading site then compute the initial estimate 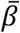 as the weighted average of all local estimates, and send it back to each site. In the second step, each site calculates and shares the local gradients 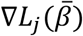 and 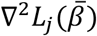. In the third step, the leading site constructs a surrogate likelihood function 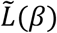 with these gradients, and obtains the surrogate estimate 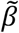.

#### Algorithm ODACH

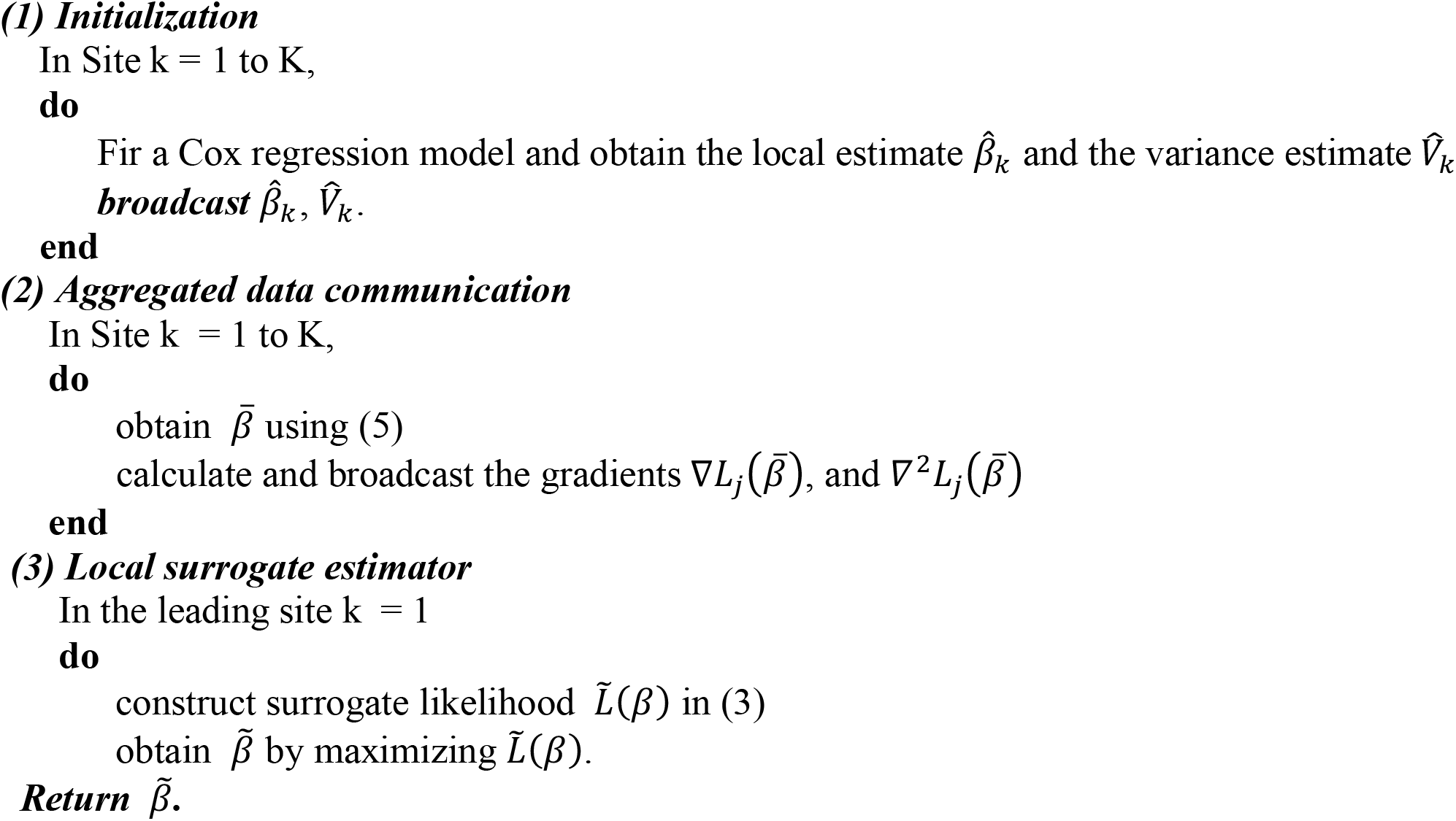

## SIMULATION STUDY

We evaluate the performance of the proposed ODAC estimator using simulated multi-site time-to-event data. A pooled dataset of N=5,000 subjects are generated based on Weibull proportional hazard model, where the baseline hazard follows a Weibull-distribution with varying scale and shape parameters. Specifically, the scale parameters range from 100 to 280 with equal space. The shape parameters range from 20 to 0.5 with equal space in the logarithm scale, see Figure 2 for an illustration. We generate 2 covariates from uniform distributions and the true log hazard ratios are set to be *β*= (−1, 1). We set the event rate (number of cases over number of subjects) as 20%, 2% and 1% by generating censoring times following appropriate distributions. The pooled data are evenly distributed to K=10 clinical sites, with 500 subjects in each site. We apply three approaches to estimate the hazard ratios of the two covariates on the time-to-event outcome, i.e. pooled stratifies Cox regression, the meta-analysis, and the proposed ODACH method. Since the pooled Cox regression estimator can be considered as a gold standard, the relative bias of meta-analysis and ODACH estimates to the pooled estimate are compared to demonstrate the advantage of ODACH. The simulation was replicated 200 times. For simplicity of illustration, we only present the results for estimation of coefficient *β*_2_ and the results for the other coefficient are similar.

**Figure 2.**
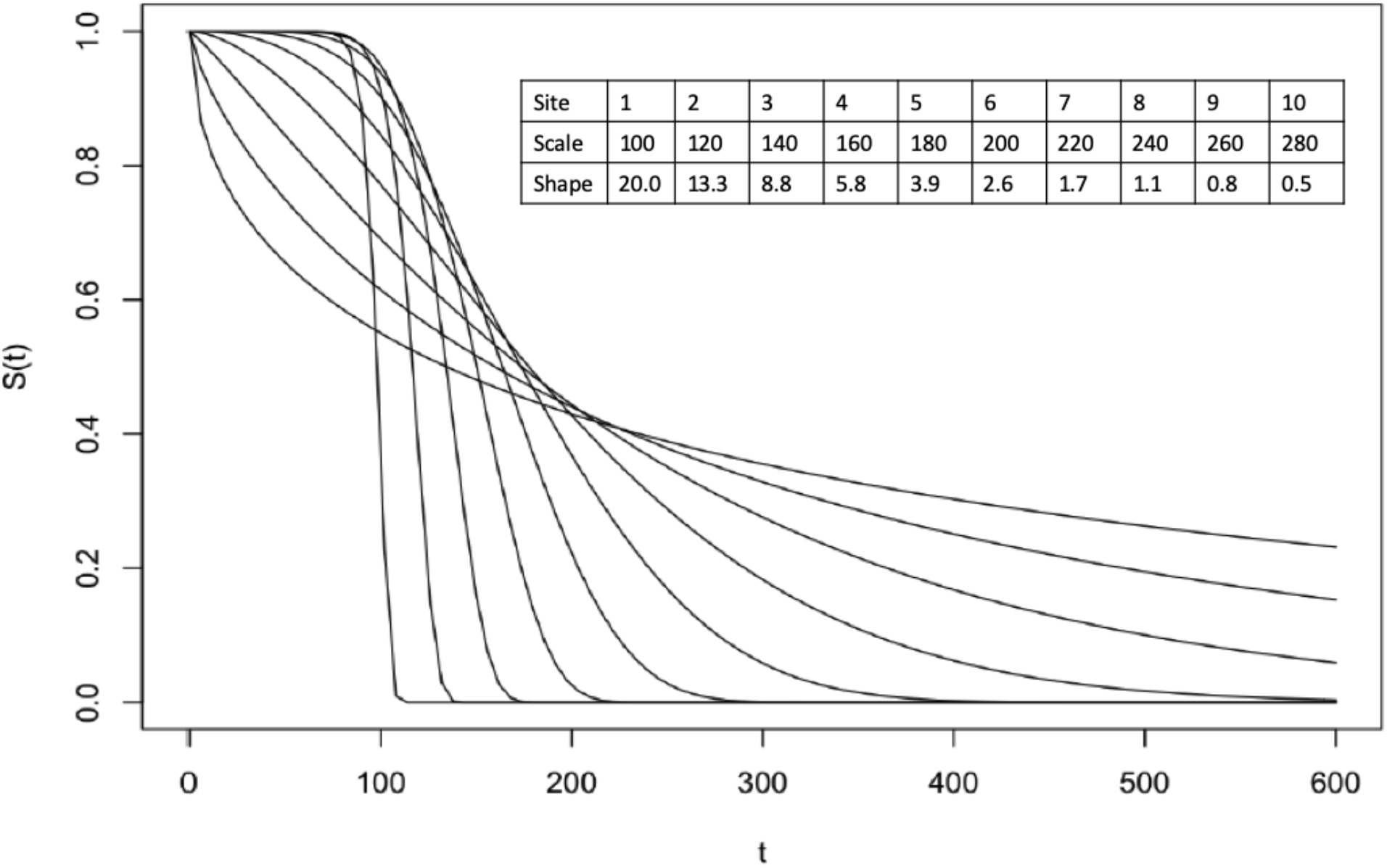
The baseline survival functions of the 10 sites in the simulated data. The varying hazard functions are Weibull functions with scale and shape parameters as listed.

The simulation results show that ODACH achieves better estimation performance than the meta-analysis estimator, especially when the event is rare. In Figure 3, ODACH obtains relative biases close to 0, meaning that it provides almost identical results to the pooled estimator, i.e. by stratified Cox model on the pooled dataset across all sites. When the event becomes more rare, meta-analysis estimator is observed to have larger bias. For example, when the event rate is 1%, the average relative bias is around -12% for the meta-analysis estimator, but only around -1% for ODACH estimator. Moreover, the variation of the meta-analysis estimator is much larger compared with that of the ODACH estimator.

**Figure 3.**
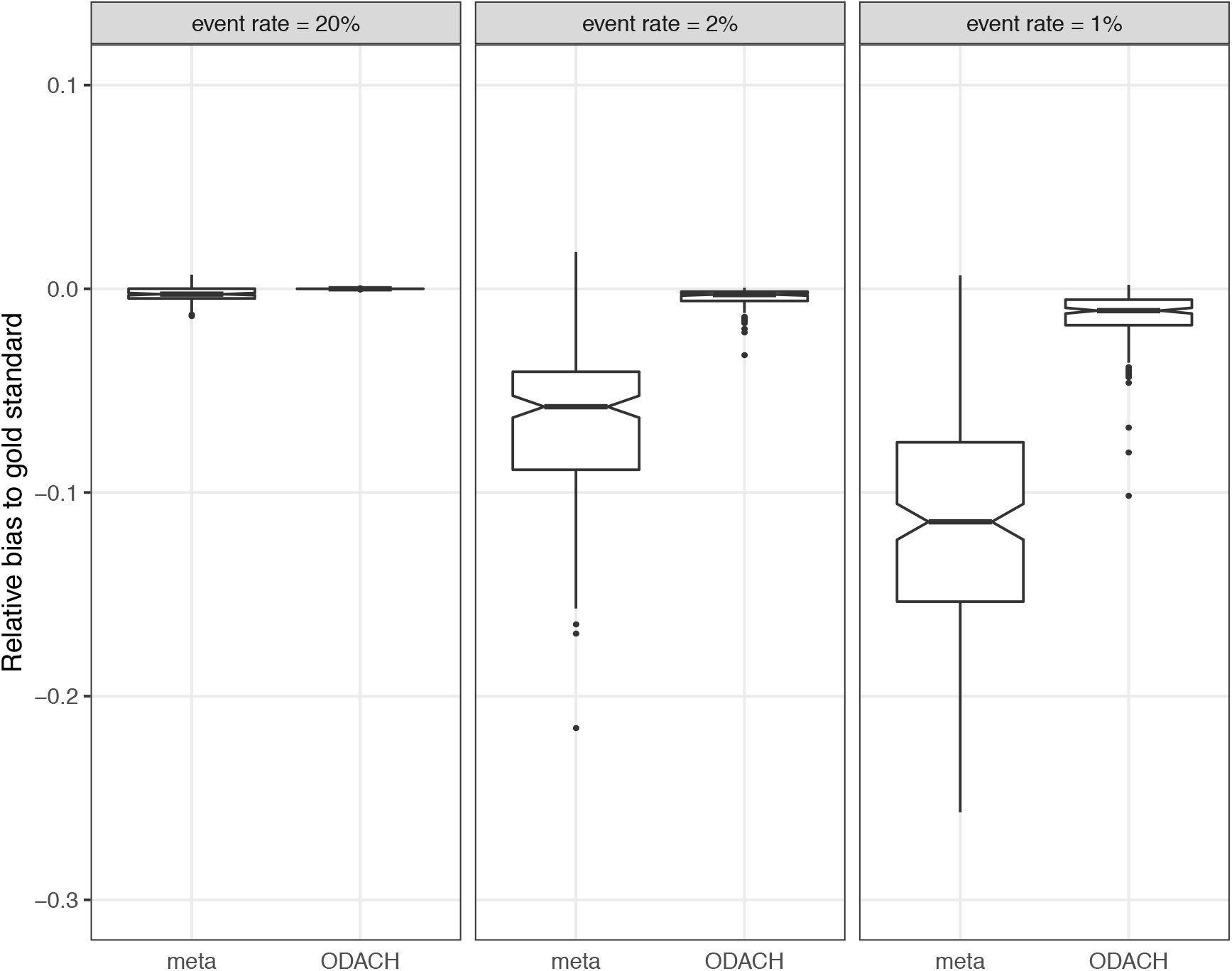
Boxplot of relative bias to the gold standard (stratified Cox model on the pooled dataset across all sites). The 2 methods compared in the plot are meta-analysis (meta) and One-shot Distributed Algorithm for Cox model with Heterogeneous baseline hazards (ODACH). The event rate varies from 20% to 1%, and under each setting, the boxplots are based on 200 replications of the simulation. The true effect size is 1.

## DISCUSSION

We developed a privacy-preserving One-shot Distributed Algorithm for the Cox model to analyze Heterogeneous multi-center time-to-event data (ODACH). The proposed surrogate likelihood approach approximates the log partial likelihood of the stratified Cox model that uses the patient-level data from all the sites. The simulation study shows the surrogate estimation obtains closer estimation towards the pooled analysis compared to the meta-analysis, especially when the event is rare. Compare to the existing One-shot Distributed Algorithm for Cox model (ODAC), ODACH allows the baseline hazard functions and covariate distributions to be site-specific, and hence is more flexible in application.

Real-world data are playing an increasing role in generating evidence to support healthcare decision making. Observational data such as EHR and medical claims data contains longitudinal information, which enables time-to-event modeling. The Cox model is one of the most commonly used models for time-to-event analysis, and has been widely applied in observational studies for treatment evaluation and risk factor identification. For example, Suchard et al. [32] conducted a systematic study to compare first-line antihypertensive drugs using a global network of administrative claims and EHR databases. The Cox model was used to study time-to-event outcomes regarding effectiveness and safety. Without sharing the patient-level data across databases, the individual estimates from each database are integrated by a meta-analysis approach. In such multi-center studies, our proposed distributed algorithm could be an alternative to the commonly used meta-analysis approach and have potential benefits in the scenario of rare events. The algorithm is implemented in the R package “pda” and is available on CRAN [33].

There are several directions of future work. Time-varying covariates or time-varying effects are sometimes encountered in time-to-event analyses [34,35]. The Cox model with these variants relaxes the proportional hazards assumption of the usual Cox model, but requires additional data for accurate estimation [36,37]. It is thus desired to develop distributed algorithm for the Cox model with time-varying covariates or time-varying effects in multi-center studies. Moreover, other survival models, such as the accelerated failure time (AFT) model [38] can be more suitable than the Cox model in certain settings. A distributed algorithm for AFT model is currently under investigation and will be reported in the future. In addition, other sources of heterogeneity other than baseline hazard functions or distributions might exist, such as missing data patterns and site-specific effect sizes. Robust methods for handling different types of heterogeneity are needed to avoid potentially misleading results.

## Data Availability

NA

## Acknowledgement

This work was supported partially through a Patient-Centered Outcomes Research Institute (PCORI) Project Program Award (ME-2019C3-18315). All statements in this report, including its findings and conclusions, are solely those of the authors and do not necessarily represent the views of the Patient-Centered Outcomes Research Institute (PCORI), its Board of Governors or Methodology Committee.

## Appendix

1. The local first order gradient 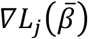

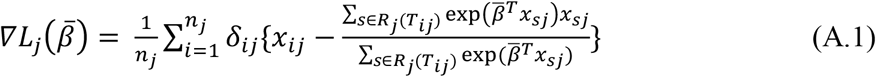
2. The local second order gradient 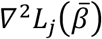

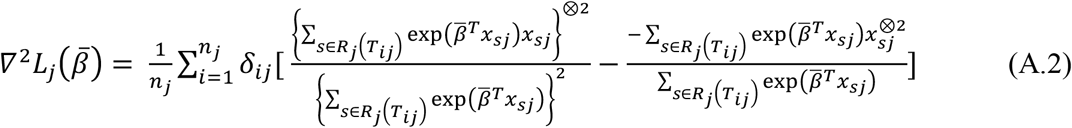

where *a*^⊗2^ for a vector *a* denotes the outer product *aa*^*T*^.
3. The global first order gradient 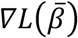

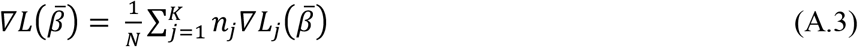
4. The global second order gradient 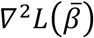

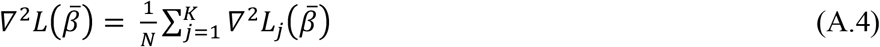

## Notes

### Competing Interest Statement

The authors have declared no competing interest.

### Author Declarations

This study does not involve human subject data.

